# Predicting the Timing of Adolescent Drinking Onset from Baseline Clinical Data: A Stacked-Encoder Survival Framework with Embedding-Derived Biotypes

**DOI:** 10.64898/2026.09.27.26364127

**Authors:** Ruobing Liu, Mohamed Azzam, Esther C. Ugwueke, Nicole L. Zabik, Shibiao Wan, Jennifer Blackford, Jieqiong Wang

## Abstract

Approximately 30% of U.S. adolescents have consumed alcohol, and earlier onset is associated with faster escalation and greater risk of adult alcohol use disorder. Predicting not only who will initiate drinking but also when could enable more timely and targeted prevention. Existing machine-learning approaches have largely classified drinking status or predefined trajectory classes, requiring fully observed outcomes and often excluding adolescents who remain abstinent at last follow-up. Many also rely on neuroimaging or small, hand-selected feature sets, limiting scalability. We developed a stacked-encoder survival framework for predicting drinking-onset timing from baseline clinical data alone. The framework compresses 1,582 clinical features using an unsupervised denoising autoencoder and re-embeds them with *FocalTab,* a TabPFN encoder fine-tuned with focal loss; the encoders are then frozen for survival modeling using a neural Cox or random survival forest head. Among 661 NCANDA adolescents who were non-drinkers at baseline, 495 initiated drinking within six years. The stacked representation outperformed single-encoder and raw-feature models, achieving an Uno C-index of 0.7654 and time-dependent AUC of 0.8274. Focal-loss encoding outperformed cross-entropy encoding, while SHAP identified alcohol expectancies, substance access, and community prevention engagement among the most influential predictors. Unsupervised clustering further identified five biotypes with a monotonic gradient in mean time to onset (4.30 to 2.86 years) and differential limbic and thalamic neuroimaging phenotypes. These findings support the feasibility of scalable, clinically based prediction of drinking-onset timing and data-driven adolescent risk stratification.

## 1. Introduction

Underage drinking refers to alcohol consumption among individuals younger than 21 years of age. According to recent reports from the Substance Abuse and Mental Health Services Administration (SAMHSA), more than 30% of U.S. adolescents have consumed alcohol at least once, and approximately 25% of those who drink report engaging in binge drinking ^1,2^. Adolescent binge drinking is associated with a range of short- and long-term adverse outcomes, including poorer academic performance, increased risk of accidents, mental health disorders, violence, liver-related complications, and an elevated risk of developing alcohol use disorder (AUD) later in life ^3^. Beyond these individual-level consequences, underage drinking also imposes substantial societal costs through increased healthcare expenditures, public safety burdens, and productivity losses ^4^. Meanwhile, earlier onset of alcohol use during adolescence is associated with more rapid escalation and a higher risk of adult alcohol use disorder (AUD) ^5^. Therefore, developing a comprehensive model to predict the onset of alcohol use among adolescents is important for identifying individuals at elevated risk and enabling timely early intervention.

Machine learning (ML) has been increasingly applied to adolescent alcohol use, primarily to predict drinking status or to distinguish drinking trajectory classes. Whelan et al. ^6^, for example, used multimodal neuroimaging together with personality, cognitive, environmental, and genetic measures to distinguish current binge drinkers from non-binge drinkers at age 14 and to predict future drinking. More recent studies have extended this framework from cross-sectional drinking status to longitudinal drinking trajectories. Zhao et al. ^7^ predicted which young adults would initiate heavy drinking after high school using mental health measures collected during adolescence. Ferariu et al. ^8^ integrated multilevel, multidomain, and multimodal imaging features from the ABCD study to distinguish youths whose alcohol sipping increased over time from those who experimented with alcohol and subsequently reduced their consumption. González et al. ^9^ developed a longitudinal network to predict monthly alcohol consumption at annual visits between ages 15 and 21, projected participants into a low-dimensional representational space, and subsequently clustered individual trajectories into six developmental pathways.

In contrast to these approaches, Nguyen-Louie et al. ^10^ modeled the timing of first drinking initiation and regular drinking onset directly in the NCANDA cohort, providing a framework more closely aligned with the onset of alcohol use. However, existing research has several limitations. First, many studies rely on neuroimaging measures ^6,8–13^ that are expensive, resource-intensive, and not readily accessible in routine or large-scale settings, which may limit their scalability and practical application. Moreover, some studies ^8,9^ have found that adding neuroimaging measures provides limited incremental value for predicting adolescent alcohol use when compared directly with models based on non-imaging measures alone. Second, predictor selection is often manual and hypothesis-driven, with features selected based on previously established risk factors ^7,9,10,14^. Although this approach facilitates hypothesis testing, it may constrain the discovery of novel predictors and interactions that are not captured by prior knowledge.

Another challenge concerns the learning regime itself. When the number of predictors approaches the number of participants and the feature space is high-dimensional, sparse, and heterogeneous, models trained directly on raw inputs are at increased risk of overfitting. Prospective studies predicting drinking onset from a single baseline assessment have generally included only several hundred participants ^7,10^. Consequently, learning a compact representation before downstream inference has become an established strategy for small-sample, high-dimensional settings. Wang et al. ^11^, for example, compressed 6,105 region-to-region connectivity measures and 16 neuropsychological measures into a joint representation because their sample of 115 participants was insufficient to support direct modeling of approximately 100,000 pairwise associations. Zhu et al. ^15^ embedded 35,778 connectivity edges into a four-dimensional nonlinear manifold.

A related challenge arises from the outcome structure in time-to-event prediction. Here, the primary imbalance is between observed events and censored observations rather than between conventional diagnostic classes. Prior studies have largely addressed this imbalance through evaluation metrics or sampling strategies rather than through the training objective itself: for example, using balanced accuracy and F1 score instead of accuracy ^7^, fitting models without explicit imbalance correction at an event rate of 6% ^16^, or relying on resampling ^8^. Focal loss ^17^ instead addresses imbalance directly within the optimization objective by assigning greater weight to difficult or underrepresented observations. In parallel, tabular foundation models ^18^ have demonstrated the potential to transfer pretrained representations to small tabular datasets, but their application to survival prediction in adolescent alcohol-use research remains unexplored.

Predicting an individual’s risk does not, by itself, capture heterogeneity in that risk. Data-driven biotyping has proven productive across psychiatric and addiction-related phenotypes. Functional-connectivity biotypes of alcohol and nicotine use disorder improved downstream diagnostic accuracy from 0.61 to 0.76 AUC ^15^; psychosis biotypes derived from multiscale connectivity cut across conventional DSM categories ^19^; and neuropsychology-informed network phenotypes mediated the association between alcohol use disorder and cognitive and motor performance ^11^. Importantly, these data-driven subtypes may provide prognostic information beyond conventional measures of disease severity. Alcohol-use severity did not differ across biopsychosocial profiles (p = 0.215), yet the profiles showed distinct three-month abstinence rates of 65.8%, 58.8%, and 36.7% ^20^.

These phenotypes are, however, derived almost exclusively in adults with established disorder ^11,15,20^ and from neuroimaging ^15,19,21,22^. The youth-focused exceptions either subtype brain regions rather than people ^22^, or subtype people without linking them to prospective substance use ^23^. Where developmental classes do exist, they are predefined rather than discovered ^8^, or describe a seven-year trajectory that can be assigned only once it has been observed ^9^. Thus, data-driven biotyping is needed to characterize heterogeneity in adolescent drinking onset and support early intervention, with subtypes validated against both the timing of drinking initiation and independent neurobiological phenotypes.

As illustrated in Figure 1, this work makes the following contributions. **First**, we predict adolescent drinking onset from **exclusively and comprehensively baseline clinical measurements** (behavioral, psychosocial, environmental, and family-history domains; 1,582 features), avoiding the cost and limited accessibility of neuroimaging as model input and enabling accessible, low-cost screening as early as possible for early intervention. **Second**, we introduce a Focal ***Stacked-Encoders* representation** that pairs an unsupervised autoencoder with a TabPFN ^18^ foundation-model encoder fine-tuned with focal loss (*FocalTab*) ^24^, leveraging a pretrained tabular model in this small-sample, high-dimensional, and censoring-imbalanced survival scenario, fused with fully connected network ^25^ CoxPH ^26^ (FCN-CoxPH) and random survival forest ^27^ (RSF) heads. **Third**, we provide interpretability and biological validation: SHapley Additive exPlanations (SHAP) ^28^ identifies the baseline predictors of earlier onset, and unsupervised **biotyping** of the learned embeddings, externally validated against independent neuroimaging phenotypes (resting-state regional homogeneity (rs-ReHo), diffusion tensor imaging (DTI) orientation-dispersion (OD), and FreeSurfer morphometry) and time-to-onset, links data-driven biotypes to limbic and prefrontal substrates of adolescent alcohol-use risk.

**Figure 1.**
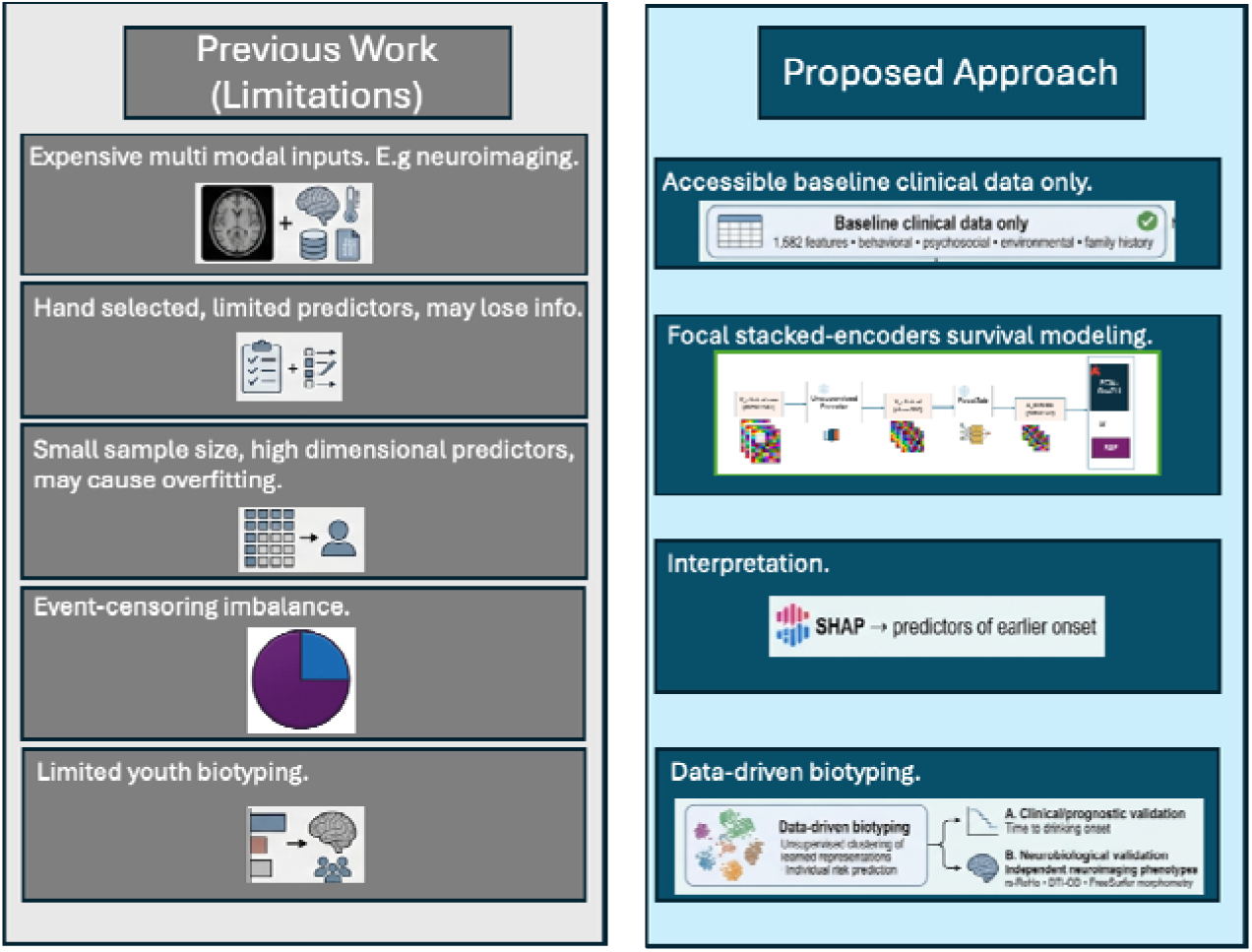
Comparison between prior adolescent alcohol-use onset approaches and the proposed framework.

## 2. Materials and Methods

### 2.1. Participants

This study used data from the National Consortium on Alcohol and Neurodevelopment in Adolescence (NCANDA), a multisite longitudinal study designed to examine the effects of alcohol exposure on adolescent neurodevelopment ^29^. Using an accelerated longitudinal design, NCANDA enrolled 831 adolescents aged 12 to 21 across five data-collection sites and conducted annual follow-up assessments. We selected 661 participants who were non-drinkers at baseline and then were followed forward; the outcome of interest was the time from the baseline assessment to the first visit at which the participant met the drinker criterion.

To be specific, drinking status was defined using the modified Cahalan inventory ^30^ applied to the Customary Drinking and Drug Use Record ^31^. After excluding participants who were drinkers at baseline or lacked follow-up assessments, the final survival cohort included 661 baseline non-drinkers (326 females and 335 males), with annual assessments conducted over 6 years. Over follow-up, 495 participants (74.9%) transitioned to drinker status (events) and 166 (25.1%) remained non-drinkers through their last available visit (right-censored).

### 2.2. Feature Preprocessing

Baseline predictors were assembled from 12 NCANDA data files, yielding an initial pool of 5,334 clinical features spanning behavioral, psychological, environmental, family, and biological domains. Features were filtered using a sequential pipeline. First, features that were completely missing across all participants were removed, leaving 4,629 features. Second, constant features with no variation across participants were excluded, leaving 4,216 features. Third, irrelevant or non-informative administrative features (e.g., identifiers, timestamps, and free-text codes) were removed, resulting in 2,718 features. Finally, features with ≥70% missing values were excluded, yielding 1,582 features for subsequent analyses. Remaining categorical items were recoded to numeric form, and residual missing values were imputed by Multivariate Imputation by Chained Equations (MICE) ^32^, fit on the training set and applied to validation and test sets to avoid leakage.

The final feature set comprised 1,582 baseline features across 23 content categories, the largest being antisocial behavior/misbehavior (533), youth behavior (227), alcohol drinking (99), sleep behavior (99), family background (94), and internalizing/psychiatric symptoms (PTSD/ADHD/depression/OCD/panic; 83).

### 2.3. Prediction of Drinker Onset

We developed a survival model to predict the time from baseline to drinker onset from baseline clinical data. Figure 2 summarizes the overall workflow, illustrating the Focal *Stacked-Encoders* representation pipeline and the survival prediction heads. Briefly, an unsupervised autoencoder compressed the high-dimensional raw clinical vector, and a TabPFN ^18^ -based encoder trained with focal loss (*FocalTab)* re-embedded it; the resulting representation was used to fit two complementary survival estimators. Section 2.3.1 defines the survival outcome; Section 2.3.2 describes the *Stacked-Encoders* pipeline; Section 2.3.3 describes the evaluation framework and ablation studies; Section 2.3.4 describes the Focal *Stacked-Encoders* model interpretation with SHAP; and Section 2.4 describes biotyping with stacked-encoder-encoded embeddings.

**Figure 2.**
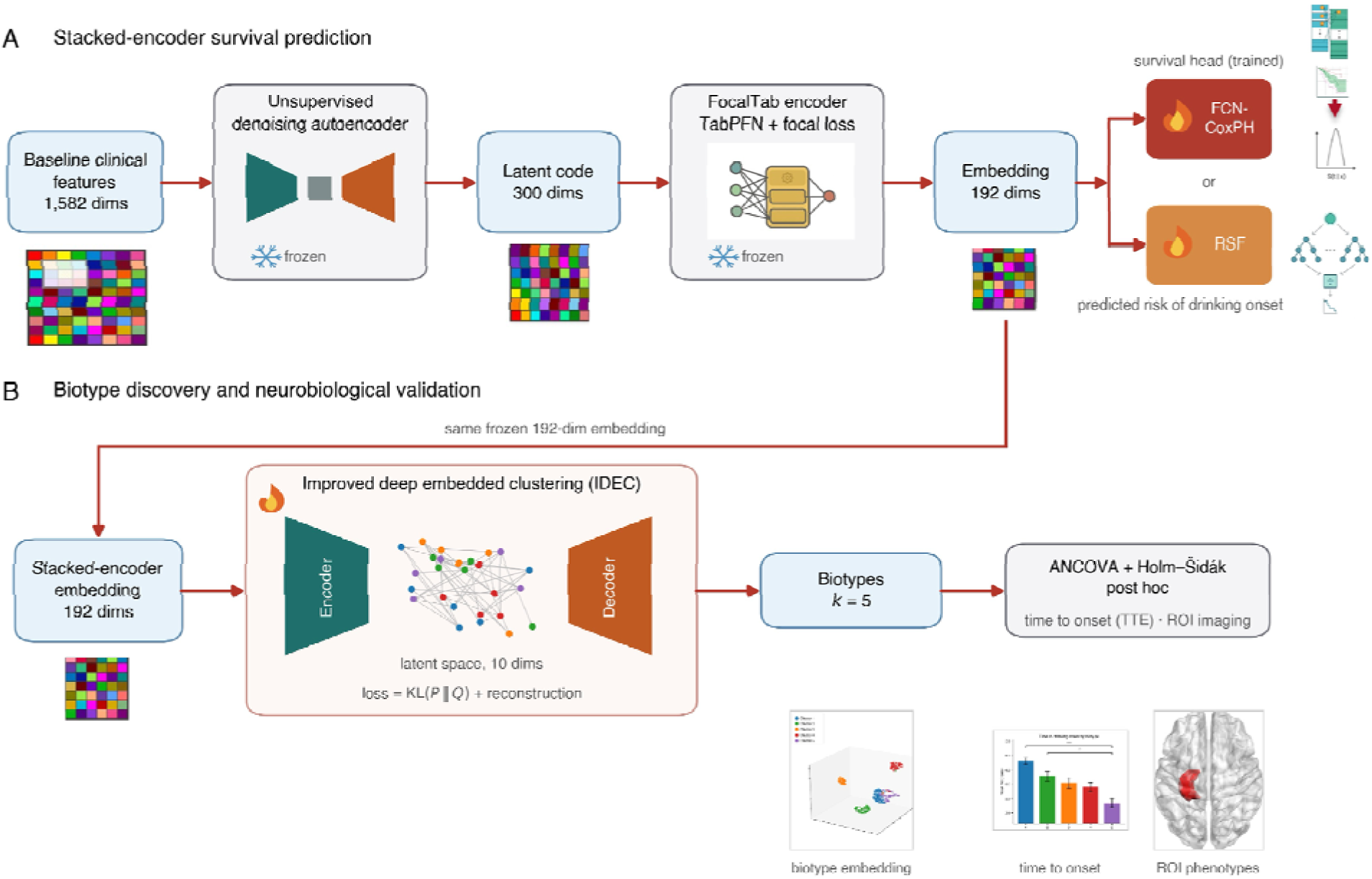
Overview of the Focal *Stacked-Encoders* framework for drinking-onset survival prediction and embedding-based biotyping. A. Focal *Stacked-Encoders* survival prediction. An unsupervised denoising autoencoder (AE) compresses the baseline clinical feature vector (1,582 dimensions) into a 300-dimensional latent code, which is re-embedded by *FocalTab*, a pretrained TabPFN tabular foundation model fine-tuned with focal loss, into a 192-dimensional representation taken from its penultimate layer. Both encoders are fit in advance and kept frozen (snowflake) during survival modeling, so that the survival outcome does not inform the learned representation. The embedding is mapped to a subject-level risk of drinking onset by one of two interchangeable trained survival heads (flame): a fully connected network optimized with the negative Cox partial log-likelihood in the CoxPH model (FCN-CoxPH), or a random survival forest (RSF) that aggregates tree-wise cumulative hazard estimates. B. Biotype discovery and neurobiological validation. The same frozen 192-dimensional Focal *Stacked-Encoders* embeddings of all 661 participants are partitioned by improved deep embedded clustering (IDEC), which jointly minimizes an autoencoder reconstruction loss and the Kullback-Leibler divergence between Student’s t soft cluster assignments and a sharpened target distribution over a 10-dimensional latent space. The resulting biotypes (k = 5) are compared on time to drinking onset (TTE) and on a priori region-of-interest (ROI) neuroimaging phenotypes by covariate-adjusted analysis of covariance (ANCOVA) with Holm-Šidák-corrected post hoc contrasts; representative outputs are shown below. Colored matrices are schematic feature matrices whose width reflects dimensionality. FCN-CoxPH, fully connected network with Cox proportional-hazards objective; IDEC, improved deep embedded clustering; ROI, region of interest; RSF, random survival forest; TTE, time to event; UMAP, uniform manifold approximation and projection.

#### 2.3.1. Definition of Drinker Onset

The survival outcome was defined on the population of baseline non-drinkers. For each participant, the event was the transition from non-drinker to drinker (moderate or heavy category by the modified Cahalan criteria ^30^), and the time-to-event (TTE) was the interval, in years, from the baseline assessment to the first follow-up visit at which the drinker criterion was met. Participants who did not meet the drinker criterion by their last available visit were treated as right-censored at that visit, resulting in 495 events and 166 censored observations.

#### 2.3.2. Focal *Stacked-Encoders* Survival Model Pipeline

The prediction pipeline comprised two stacked representation-learning encoders followed by a survival head (Figure 2A). An unsupervised denoising autoencoder first compressed and denoised the 1,582-dimensional clinical vector into a compact embedding; a TabPFN-based encoder fine-tuned with focal loss (*FocalTab*) then re-embedded this representation using a pretrained tabular prior; and a survival estimator mapped the resulting embedding to a predicted risk of drinker onset. Both encoders were fit before and held fixed (frozen) during survival-head training, so that no information from the survival outcome could leak into the learned representation.

##### 2.3.2.1. Unsupervised Denoising Autoencoder (AE)

To learn a robust low-dimensional representation of the sparse, heterogeneous clinical space, we trained an unsupervised denoising autoencoder (shortened as AE) on the standardized features. During training, the input was corrupted by randomly masking (zeroing) a random subset of features, and the network was required to reconstruct the uncorrupted input; this denoising objective discourages reliance on any individual feature and yields noise-robust embeddings. The encoder mapped the input to a bottleneck embedding Z through a stack of Linear -> LayerNorm -> GELU -> Dropout -> Linear layers, and a single linear decoder reconstructed the input (X^) from Z. The training objective was the mean-squared reconstruction error between X and X^ together with an L2 penalty on the embedding norm (with L2 weight decay on the parameters), which regularizes the latent geometry and discourages degenerate solutions. To prevent leakage, the autoencoder was fit on the training split only and then applied unchanged to the validation and test splits. The bottleneck width was treated as a hyperparameter and swept from 100 to 500 dimensions; for each candidate width, a Cox proportional-hazards model was fit on the resulting embedding and scored by the validation concordance index (C-index), and the best-performing width (300 dimensions) was retained for all subsequent experiments.

##### 2.3.2.2. FocalTab Encoder (FocalTab)

The second encoder transferred a pretrained tabular prior to the small-sample survival problem. TabPFN ^18^ is a pretrained, transformer-based foundation model for tabular data that approximates Bayesian inference over a synthetic prior via alternating row- and column-wise attention in a single forward pass. We adapted TabPFN as a supervised feature extractor by fine-tuning it with focal loss ^17^, on the auxiliary binary task of classifying the final event as drinkers versus non-drinkers; focal loss down-weights easily classified majority examples and concentrates learning on the minority (non-drinker) class, mitigating the class imbalance. Embeddings were extracted with a strictly split-respecting, three-phase protocol. In Phase A (train -> val), standardization and TabPFN fine-tuning were fit on the training split only across a learning-rate grid, and the validation split was used solely to select the number of epochs and the learning rate, the focal-loss parameters α and γ (by validation AUC), and the operating probability threshold (by Youden’s J statistic). In Phase B (train+val -> test), standardization was refit on the combined training and validation data, and TabPFN was refit on train+val using exactly the epoch, learning rate, and focal-loss parameters selected in Phase A. In Phase C, the model was evaluated in a single pass on the held-out test split using the Phase A threshold. For every subject in each split, the model’s penultimate (pre-classification) representation, a 192-dimensional embedding, was extracted and saved as the *FocalTab* representation. As with the autoencoder, the *FocalTab* encoder was frozen when the survival head was trained.

##### 2.3.2.3 Survival Heads

The frozen embedding was mapped to a survival prediction by one of two complementary estimators. FCN ^25^-CoxPH ^26^ is a fully connected neural network that outputs a single continuous log-relative-risk score and is trained by minimizing the negative Cox proportional-hazards partial log-likelihood; its tunable hyperparameters were the learning rate, weight decay, dropout rate, and the number and width of the fully connected layers. The Random Survival Forest ^27^ (RSF) is an ensemble of survival trees grown with the log-rank splitting rule that aggregates tree-wise Nelson-Aalen estimates into a subject-specific cumulative hazard function, providing a nonlinear, proportional-hazards-free complement to the Cox model; its tunable hyperparameters were the minimum number of samples required to split a node, the minimum number of samples per leaf, and the maximum tree depth.

Models were optimized with a multi-stage protocol that rigorously segregated training, validation, and test data (Figure 2A). In Phase A (training and hyperparameter selection), feature selection and feature scaling were fit on the training set only; the survival head was trained on the training set, and Harrell’s C-index was scored on the validation set after every epoch. The weights from the best validation epoch were retained, and training was stopped when validation C did not improve for a fixed patience; the selected epoch was recorded as “chosen_epoch”. In Phase B (refitting), feature selection and the scaler were refitted on the combined training + validation cohort (TRVAL), and a fresh model was trained from scratch on TRVAL for exactly “chosen_epoch” epochs, so that the final model used the maximum available development data while inheriting the early-stopping decision made on held-out data. In Phase C (test), the refitted model was evaluated once on the strictly held-out test set, with all configurations fixed from the validation phase.

Hyperparameters were tuned on the validation set by grid search. For FCN-CoxPH, the tuned hyperparameters were the learning rate, weight decay, dropout, and the number of fully connected layers; for RSF, they were the minimum samples per split, minimum samples per leaf, and maximum tree depth. The baseline and ablation protocols followed an identical three-phase procedure, which is described in Section 2.3.3.

#### 2.3.3. Evaluation and Ablation Studies

Model performance was evaluated along four complementary survival metrics: Harrell’s C-index and the Uno C-index (discrimination, with the Uno index inverse-probability-of-censoring weighted to correct for censoring bias), the Integrated Brier Score (IBS) (calibration and overall accuracy, lower is better), and the time-dependent AUC (tdAUC). 95% confidence intervals (CI) were obtained by bootstrap resampling of the test set.

We conducted an ablation over representation configurations to isolate the contribution of each encoder. Six input representations were compared, each with both survival heads (FCN-CoxPH and RSF): (1) **Clinical raw** (the 1,582-dimensional features directly); (2) **unsupervised encoder only**; (3) ***FocalTab* encoder only**; (4) **cross-entropy TabPFN encoder only** (the original same TabPFN encoder trained with standard cross-entropy rather than focal loss, shortened as CE-TabPFN); (5) **AE + *FocalTab*** (the Focal *Stacked-Encoders* pipeline); and (6) **AE + CE-TabPFN** (the CE *Stacked-Encoders* pipeline). Comparing configurations (5) and (6) isolated the effect of the focal-loss encoder, and comparing (2) (3) (4) against (5) (6) isolated the benefit of stacking the two encoders.

#### 2.3.4. Subgroup Evaluation by Sex

To assess whether predictive performance generalized across sex, the best-performing configuration (AE + *FocalTab*, FCN-CoxPH head) was additionally evaluated separately in female and male test participants. The predictions from the single held-out test-set evaluation described in Section 2.3.3 were stratified by sex, and Uno’s and Harrell’s C-index, IBS, and the tdAUC were recomputed within each subgroup, with 95% CIs obtained by bootstrap resampling within that subgroup. Because each subgroup is small (44 female, 56 male), subgroup estimates are reported descriptively.

#### 2.3.5. Model Interpretation with SHAP

SHapley Additive exPlanations (SHAP) ^28^ were used to identify the contribution of individual baseline clinical features to the predicted risk of drinker onset within the best-performing configuration (AE + *FocalTab*, FCN-CoxPH head). SHAP values were computed over the clinical feature space, and features were ranked by mean absolute SHAP value to identify the most influential predictors (Figure 5). A beeswarm summary was used to display, for the top-ranked features, both the magnitude and the direction of each feature’s effect on predicted risk.

### 2.4. Biotyping with Focal *Stacked-Encoders* Embeddings

#### 2.4.1 Biotype Discovery

To examine whether the learned representation captured clinically and biologically meaningful heterogeneity, we performed unsupervised biotyping on the Focal *Stacked-Encoders* embeddings. Clustering operated on the 192-dimensional representation produced by the frozen AE and *FocalTab* encoders (Section 2.3.2); the clustering pooled all 661 participants. Because clustering is unsupervised and no outcome is predicted, the pooled embeddings were first standardized (each of the 192 dimensions z-scored). Biotypes were identified with Improved Deep Embedded Clustering (IDEC), which jointly learns a low-dimensional embedding and cluster assignments. A stacked autoencoder was pre-trained by reconstruction (mean-squared error; 200 epochs; Adam, learning rate 0.001), and cluster centers were initialized by K-means (k = 2-9, 20 initializations) on the 10-dimensional latent space. The encoder and cluster centers were then jointly optimized to minimize the Kullback-Leibler divergence between a Student-t soft-assignment distribution (Q) and a sharpened target distribution (P, recomputed every 20 iterations), regularized by the reconstruction loss (IDEC weight gamma = 0.10), for up to 400 full-batch iterations, stopping when fewer than 0.1% of assignments changed between updates. Each participant was assigned to the biotype with the highest soft-assignment probability.

For the unsupervised clustering, model selection and validation targeted robustness rather than predictive accuracy. The number of clusters k was chosen by consensus across internal quality indices (silhouette, Calinski-Harabasz, and Davies-Bouldin), and the within-cluster sum-of-squares elbow; agreement (ARI) with the K-means initialization and the separation of survival curves across clusters (log-rank p) were additionally examined. Formal hypothesis testing was reserved for the downstream neuroimaging and time-to-event comparisons described below.

#### 2.4.2. Biotype TTE Associations

Differences in time to drinking onset (TTE) across biotypes were tested by analysis of covariance (ANCOVA) on the observed TTE, with biotypes and baseline age, sex, and site as covariates. The omnibus biotype effect was evaluated with a Type-II sum-of-squares *F* test and its effect size reported as partial *η*^2^. Pairwise differences between biotypes were then examined with covariate-adjusted contrasts, correcting the ten comparisons with the Holm-Šidák procedure.

#### 2.4.3. Biotype Neuroimaging Associations

The five biotypes were externally validated against independent neuroimaging phenotypes derived from the NCANDA multimodal diffusion Magnetic Resonance Imaging (MRI) resting-state function, diffusion microstructure, and structural morphometry (preprocessing described in Section 2.5). Guided by the neurocircuitry of adolescent reward, emotion, and alcohol-use risk, analyses were restricted to an a priori set of bilateral regions of interest spanning the amygdala, anterior hippocampus, hypothalamus, anterior insula (the frontoperculo-insular salience/ventral-attention region), ventromedial prefrontal cortex, and anterior thalamus, specifically the left and right amygdala, hippocampus, hypothalamus, salience-network frontoperculo-insula, thalamus), and suborbital/ventromedial prefrontal cortex, defined within the corresponding structural (aseg, Destrieux aparc.a2009s, and wmparc), functional, and diffusion parcellations. For each imaging measure, an ANCOVA tested the main effect of biotype (k = 5) on each region of interest.

Measures and nuisance covariates were modality-specific. For MRI, we tested fractional anisotropy (FA), radial, mean, and axial diffusivity (RD, MD, AD), the NODDI intracellular and isotropic volume fractions (ICVF, ISOVF), and the orientation dispersion index (OD), adjusting for age, sex, and site. For resting-state functional magnetic resonance imaging (rs-fMRI), we tested regional homogeneity (ReHo), the amplitude of low-frequency fluctuations (ALFF), and functional connectivity (FC), adjusting for age, sex, site, and mean framewise displacement (mean FD). For structural MRI, we tested average cortical thickness, surface area, and gray-matter volume, adjusting for age, sex, site, and intracranial volume (ICV).

Within each modality, omnibus biotype effects were corrected across regions and measures using the Benjamini-Hochberg false discovery rate (FDR). After FDR correction, a single measure survived per functional and diffusion modality, ReHo for resting-state function and the OD for diffusion, whereas structural morphometry reached only nominal significance. For regions with a significant omnibus biotype effect, pairwise differences between biotypes were examined post hoc with Holm-Šidák correction. Differences in time-to-event across biotypes were tested analogously by ANCOVA on TTE, adjusting for age, sex, and site, with Holm-Šidák-corrected pairwise contrasts.

### 2.5. Neuroimaging Preprocessing

All imaging data were organized in Brain Imaging Data Structure ^33^ (BIDS) format. The detailed protocol of NCANDA MRI, rs-fMRI, and DTI data acquisition can be found in the NCANDA manual ^29^.

#### 2.5.1. Structural MRI (FreeSurfer)

T1-weighted images were processed with fMRIPrep ^34^ 24.1.1, which performed intensity non-uniformity correction, skull-stripping, tissue segmentation, cortical-surface reconstruction with FreeSurfer recon-all, and spatial normalization to MNI152NLin2009cAsym via ANTs ^35^. Cortical surfaces were parcellated with the Destrieux ^36^ (aparc.a2009s) atlas to yield regional cortical thickness (e.g., the suborbital/ventromedial prefrontal cortex), and intracranial volume was retained as a covariate.

#### 2.5.2. Resting-state fMRI (ReHo)

Functional images were preprocessed with fMRIPrep 24.1.1 (head-motion correction, susceptibility-distortion correction, boundary-based coregistration to the T1-weighted reference, and normalization to MNI152NLin2009cAsym and MNI152NLin6Asym) and post-processed with XCP-D ^37^ 0.10.5. After discarding 10 initial volumes, a 27-parameter confound model was regressed out; the time series were band-pass filtered (0.01-0.08 Hz) and smoothed (6 mm FWHM), and regional homogeneity (ReHo), the Kendall coefficient of concordance between each voxel’s time series and that of its neighboring voxels, was computed and summarized over the Schaefer-based 4S156 atlases ^38–41^ (e.g., hippocampus, amygdala).

#### 2.5.3. Diffusion MRI (NODDI orientation dispersion)

Diffusion-weighted images were preprocessed with QSIPrep ^42^ 1.0.0: MP-PCA denoising ^43^ (dwidenoise), Gibbs de-ringing (mrdegibbs), B1 bias-field correction, FSL ^44^ eddy head-motion and eddy-current correction, TOPUP susceptibility-distortion correction, and rigid coregistration to the T1-weighted reference (output resolution 1.875 mm for GE and 2.5 mm for Siemens). Tissue microstructure was then reconstructed with QSIRecon 1.0.0 using the AMICO implementation of the NODDI ^45^ model, yielding the OD, which was summarized over the AAL116 and 4S156 atlases.

## 3. Results

### 3.1. Demographics Analysis

In Table 1, the analytic cohort comprised 661 baseline non-drinkers (326 female, 335 male; median baseline age within the 12-21 years enrollment range). Over the six-year follow-up, 495 participants (74.9%) transitioned to drinker status and 166 (25.1%) were right-censored, with time-to-onset ranging from 1 to 6 years (median 4). Consistent with normative adolescent development, baseline internalizing symptom scales were low and floored across informants, whereas alcohol-expectancy scales showed substantial variation; cohort characteristics are summarized in Table 1. For model development, participants were partitioned into training (n = 462), validation (n = 99), and test (n = 100) sets (approximately 7:1:2), while maintaining similar event/censored rates.

**Table 1.** Demographic characteristics of non-drinkers and drinkers group.

|  | **Train** | **Val** | **Test** | **Overall** |
| --- | --- | --- | --- | --- |
| **N subjects** | 462 (69.9%) | 99 (15.0%) | 100 (15.1%) | 661 (100%) |
| **Sex** |  |  |  |  |
| Female | 228 (49.4%) | 54 (54.5%) | 44 (44.0%) | 326 (49.3%) |
| Male | 234 (50.6%) | 45 (45.5%) | 56 (56.0%) | 335 (50.7%) |
| **Age at baseline (yrs)** | 15.75 ± 2.33 | 15.57 ± 2.28 | 15.28 ± 2.25 | 15.65 ± 2.31 |
| **Race** |  |  |  |  |
| White/Caucasian | 326 (70.6%) | 66 (66.7%) | 76 (76.0%) | 468 (70.8%) |
| Black/African-American | 54 (11.7%) | 12 (12.1%) | 12 (12.0%) | 78 (11.8%) |
| Asian | 42 (9.1%) | 5 (5.1%) | 3 (3.0%) | 50 (7.6%) |
| Native American | 1 (0.2%) | 2 (2.0%) | 0 (0.0%) | 3 (0.5%) |
| Pacific Islander | 2 (0.4%) | 1 (1.0%) | 0 (0.0%) | 3 (0.5%) |
| Hispanic | 52 (11.3%) | 12 (12.1%) | 12 (12.0%) | 76 (11.5%) |
| Other | 37 (8.0%) | 13 (13.1%) | 9 (9.0%) | 59 (8.9%) |
| **Outcome: drinker onset** |  |  |  |  |
| Events | 346 (74.9%) | 74 (74.7%) | 75 (75.0%) | 495 (74.9%) |
| Censored | 116 (25.1%) | 25 (25.3%) | 25 (25.0%) | 166 (25.1%) |

### 3.2. Comparison with Other Methods and Ablation Studies

Table 2 reports test-set survival performance for six input representations: the raw clinical features, each encoder alone (AE, *FocalTab*, and CE-TabPFN encoder), and the two *Stacked-Encoders* (AE + FocalTab, AE + CE-TabPFN), each evaluated with the FCN-CoxPH and RSF heads using Uno’s and Harrell’s C-indexes (shortened as Uno C and Harrell C), IBS, and tdAUC, with 95% bootstrap CIs (Table 2, Figure 3). Across both heads, the Focal *Stacked-Encoders* pipeline showed the strongest overall performance across discrimination, calibration, and tdAUC metrics. The RSF variant achieved the highest Uno C of 0.7654 and the highest tdAUC of 0.8274, while the FCN-CoxPH variant yielded the lowest IBS (0.1708) and the highest Harrell C (0.7557).

**Table 2.**
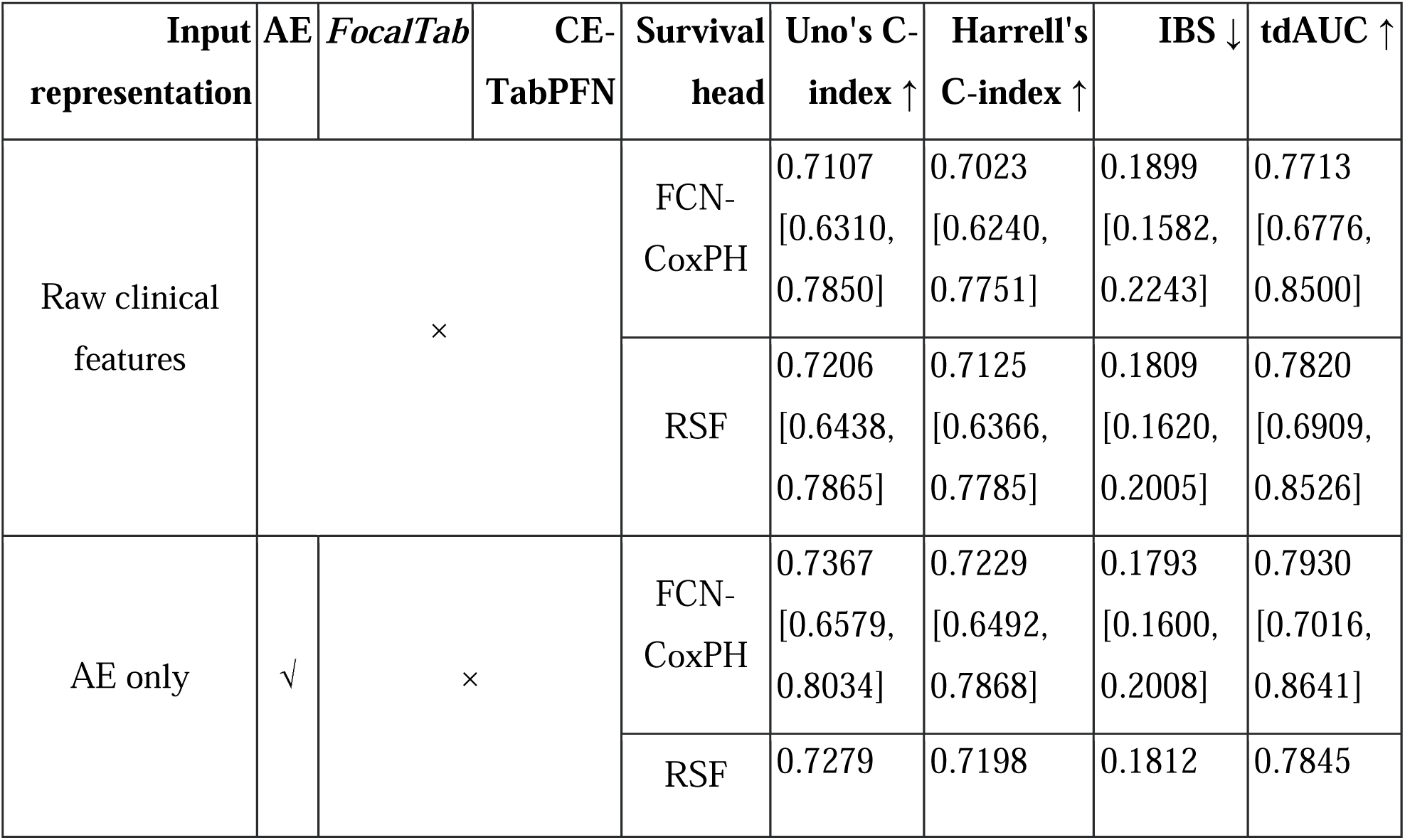
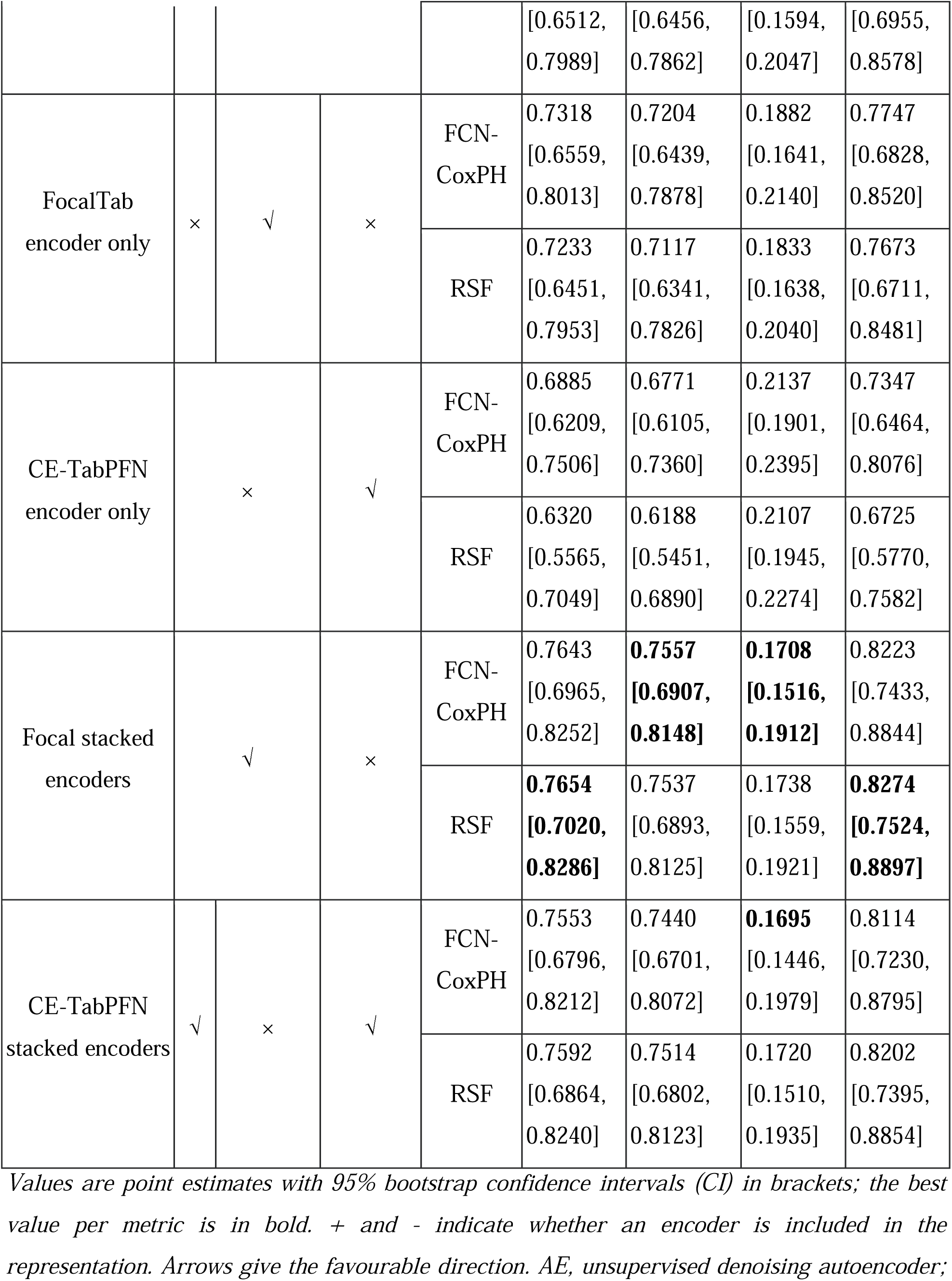
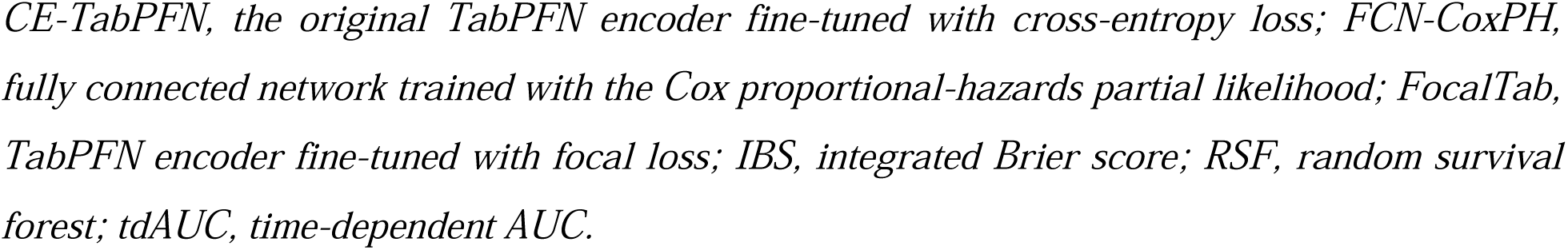
Test-set survival performance by representation configuration and survival head (95% bootstrap CI).

**Figure 3.**
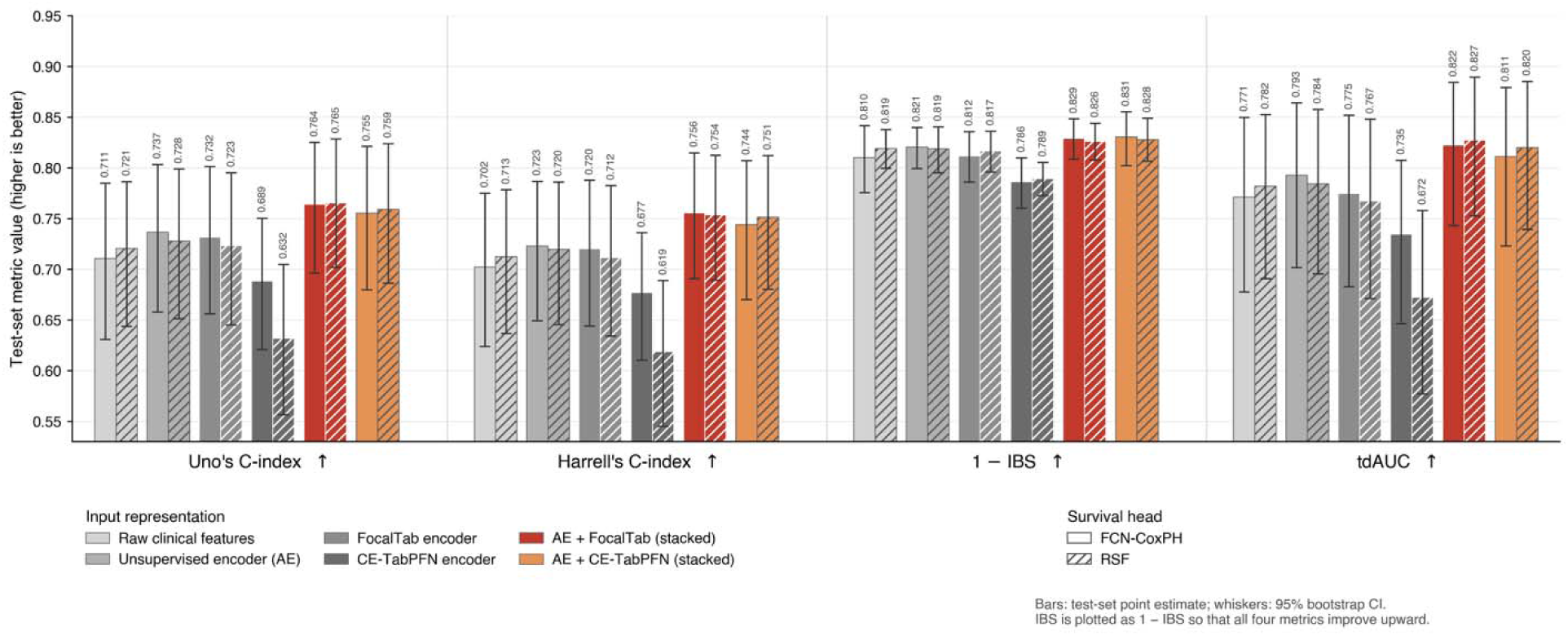
Survival performance across input representations and survival heads. Six input representations: the raw clinical features (1,582 dimensions), each encoder alone (AE, the *FocalTab* encoder, and the CE-TabPFN encoder), and the two stacked combinations were each evaluated with both survival heads on the held-out test set. Bar color encodes the input representation and hatching the survival head (solid, FCN-CoxPH; hatched, RSF). Bars are point estimates, and whiskers are 95% bootstrap confidence intervals (CIs); the value above each bar is the point estimate. Metrics are grouped along the x-axis, with arrows giving the favorable direction: Uno’s and Harrell’s concordance indices and the time-dependent AUC increase with better performance, and the IBS is plotted as 1 - IBS so that all four metrics improve upward. Across both heads, the two stacked representations exceeded every single-encoder and raw-feature configuration on discrimination and time-dependent accuracy, and the CE-TabPFN encoder alone was the weakest configuration overall. Exact values with confidence intervals are given in Table 2. AE, unsupervised denoising autoencoder; CE-TabPFN, TabPFN encoder fine-tuned with cross-entropy loss; FCN-CoxPH, fully connected network trained with the Cox proportional-hazards partial likelihood; *FocalTab*, TabPFN encoder fine-tuned with focal loss; IBS, integrated Brier score; RSF, random survival forest; tdAUC, time-dependent AUC.

#### 3.2.1. Stacked Representation Outperforms Single and Raw Encoders

The two *Stacked-Encoders* configurations outperformed every single-encoder and raw-feature configuration on all four metrics. Relative to the raw-feature baseline (RSF: Uno C = 0.7206, tdAUC = 0.7820, IBS = 0.1809), the Focal *Stacked-Encoders* representation improved discrimination and accuracy by about 0.045 (RSF: Uno C = 0.7654, tdAUC = 0.8274) while lowering the IBS (0.1708-0.1738). Each encoder was also beneficial on its own: the AE (FCN-CoxPH: Uno C = 0.7367, tdAUC = 0.7930) and FocalTab (FCN-CoxPH: Uno C = 0.7318, tdAUC = 0.7747) each exceeded the raw baseline (FCN-CoxPH: Uno C = 0.7107, tdAUC = 0.7713), but neither matched the stacked model, indicating that the denoising compression and the focal-loss tabular re-embedding contribute complementary information that is best exploited jointly. This ordering (stacked > single encoder > raw features) held for both the FCN-CoxPH and RSF heads.

#### 3.2.2. Focal-Loss Encoding Outperforms Cross-Entropy under Class and Censoring Imbalance

Because events of non-drinkers were a minority and the time-to-event target is heavily event-dominated, the objective used to train the TabPFN encoder mattered substantially. Encoding with a standard cross-entropy TabPFN, the off-the-shelf objective, was the weakest configuration overall, falling below even the raw features (FCN-CoxPH: Uno C = 0.6885, IBS = 0.2137; RSF: Uno C = 0.6320, tdAUC = 0.6725), whereas the focal-loss FocalTab encoder recovered and surpassed the raw baseline (FCN-CoxPH: Uno C = 0.7318). The advantage persisted after stacking on the autoencoder: FocalTab *Stacked-Encoders* exceeded CE-TabPFN *Stacked-Encoders* on discrimination and accuracy for both heads (RSF: Uno C = 0.7654 vs. 0.7592, tdAUC = 0.8274 vs. 0.8202; FCN-CoxPH: Uno C = 0.7643 vs. 0.7553), although the autoencoder narrowed the gap by partially compensating for the weaker cross-entropy embedding. Focusing the encoder’s learning on the hard, minority (non-drinker) cases through focal loss therefore yielded representations that transferred better to the imbalanced survival problem than the default CE-TabPFN.

#### 3.2.3. Comparison of Survival Heads (FCN-CoxPH versus RSF)

The two survival heads performed comparably and benefited similarly from the learned representations. For the best (Focal stacked encoders) configuration, RSF gave marginally higher discrimination and accuracy (Uno C = 0.7654, tdAUC = 0.8274) while FCN-CoxPH gave marginally better calibration and rank agreement (IBS = 0.1708, Harrell C = 0.7557); the bootstrap CIs overlapped substantially, so the two heads were effectively equivalent on the strongest representation. RSF was, however, more sensitive to uninformative inputs: on the CE-TabPFN encoding, its discrimination fell further (Uno C = 0.6320) than FCN-CoxPH’s (0.6885), consistent with tree ensembles being more affected by noisy features. Overall, model performance was more strongly influenced by the choice of representation than by the choice of survival head.

### 3.3. Subgroup Evaluation by Sex

Performance was comparable in female and male participants (Table 3, Figure 4). For the best configuration (AE + *FocalTab,* with the FCN-CoxPH head), the Uno C-index was 0.7374 [0.6234, 0.8355] in females (35 events / 44) and 0.7798 [0.6848, 0.8680] in males (40 events / 56), with tdAUC essentially identical (0.8181 vs. 0.8183) IBS of 0.1739 and 0.1683. The male group performed slightly better than the female group.

**Table 3.**
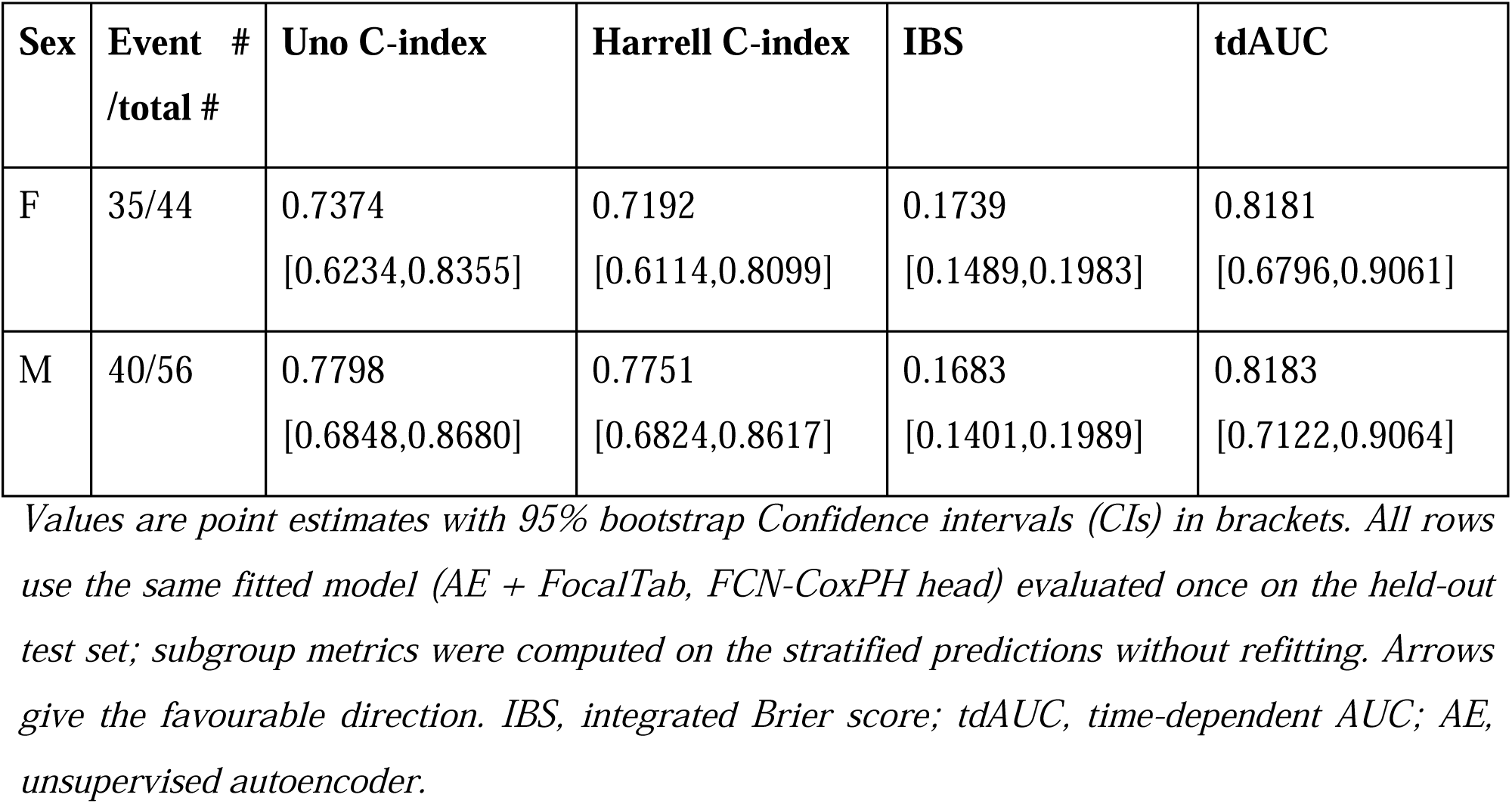
Survival performance of the final model by sex.

**Figure 4.**
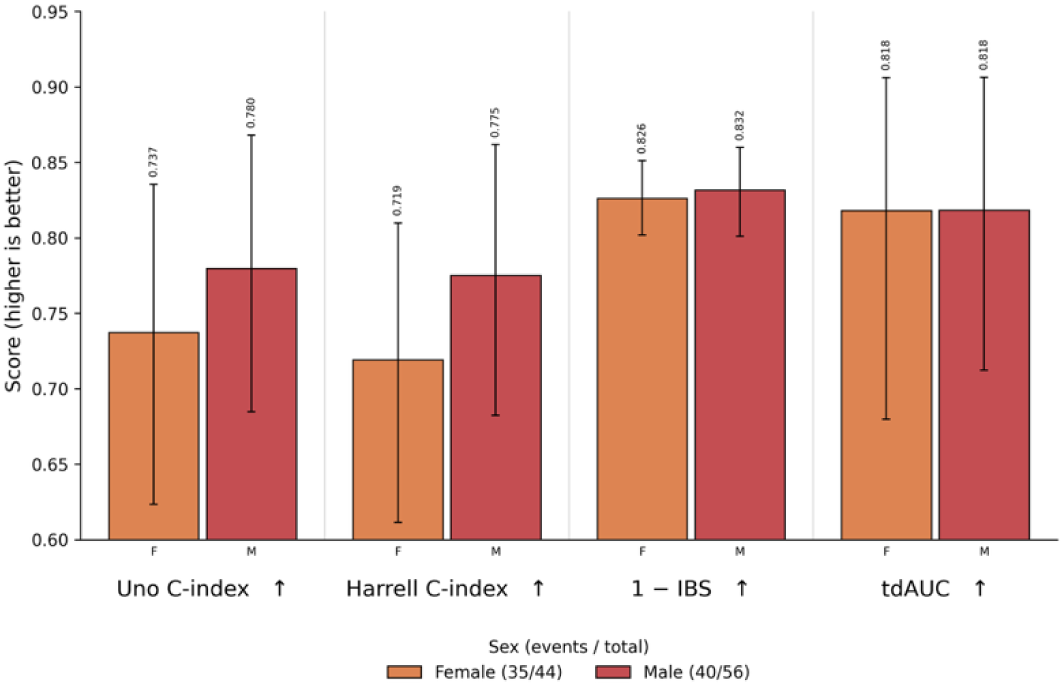
Performance of the best configuration by sex. The AE + *FocalTab* representation with the FCN-CoxPH head, evaluated separately in female (35 events / 44) and male (40 events / 56) test participants; subgroup intervals were obtained by bootstrap resampling within each subgroup. Confidence intervals (CIs) overlap substantially in every metric. The IBS is plotted as 1 - IBS so that all four metrics improve upward, as indicated by the arrows. IBS, integrated Brier score; tdAUC, time-dependent AUC.

### 3.4. Model Interpretation with SHAP

Figure 5 summarizes the SHAP interpretation of the best model (AE + FocalTab, FCN-CoxPH), showing both the per-subject impact of each feature on predicted risk (beeswarm, left) and the mean absolute SHAP value on overall feature importance (bar plot, right). The highest-ranked predictors formed four coherent themes. The single most influential features concerned substance access and procurement: from whom and where drugs had most recently been obtained, and how difficult alcohol or drugs were to obtain, including at home. A large, contiguous block of alcohol-expectancy (AEQ) items followed, dominated by positive social and affective expectancies: that drinking makes parties more fun, makes it easier to be with others and the world seem nicer, relaxes people, makes the future seem brighter, makes one feel stronger and more powerful, and makes it easier to speak in front of a group or to join in with others. Neighborhood and community-engagement items, residents’ influence on local policy, their efforts to prevent teen drinking, and their participation in local activities, together with a circadian/behavioral marker (wake-up time), rounded out the leading predictors.

**Figure 5.**
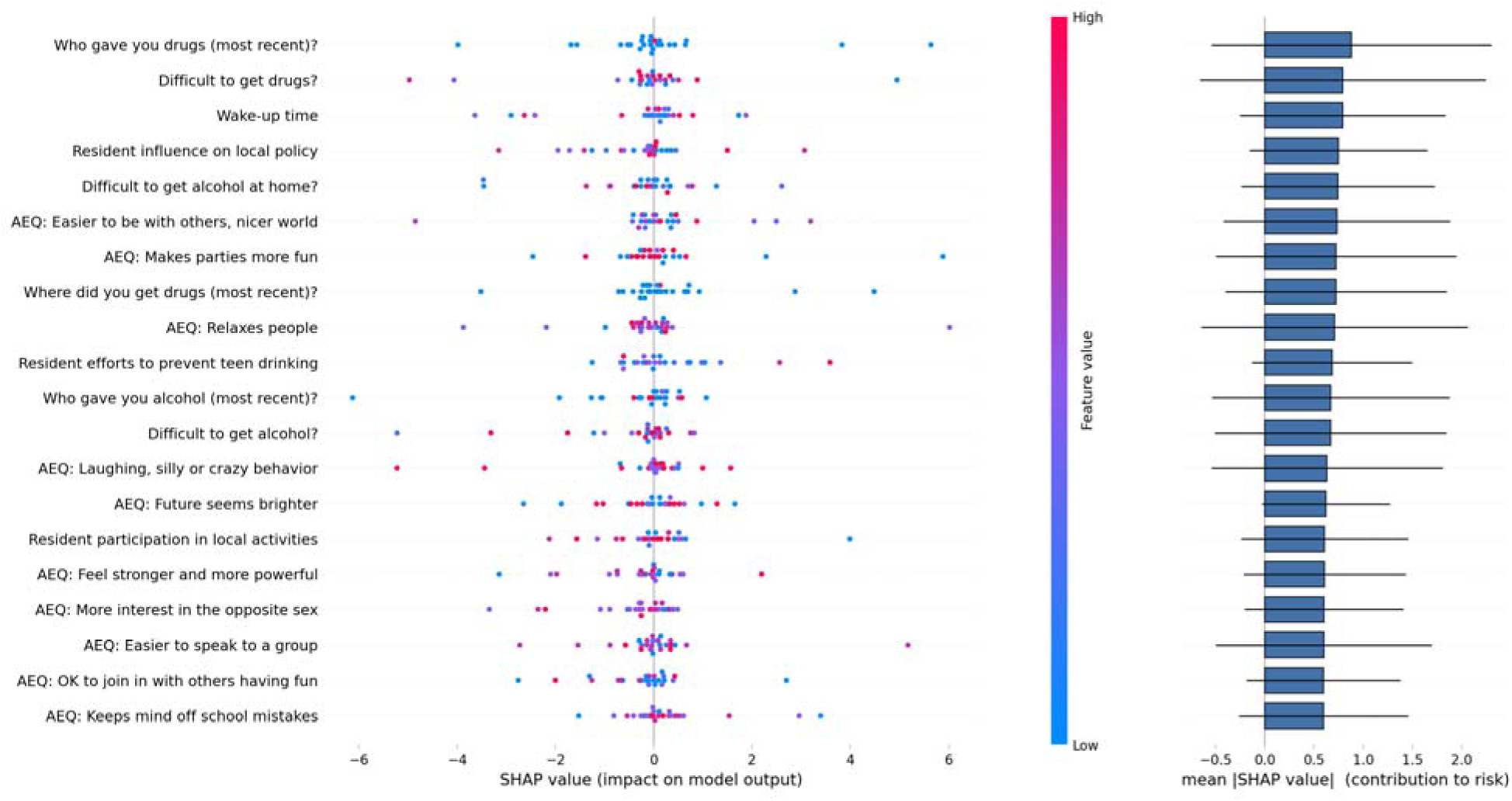
SHAP interpretation of the most important clinical features: per-subject contributions (beeswarm, colored by feature value, left) and mean absolute SHAP value (overall importance, right).

The beeswarm indicated a consistent direction for the expectancy items: stronger endorsement (red) shifted predictions toward higher risk (positive SHAP, earlier onset), whereas weaker endorsement (blue) was protective, reinforcing alcohol expectancy as a modifiable risk signal. Easier access to alcohol or drugs and lower perceived community engagement in prevention likewise tended to increase predicted risk. What adolescents expected alcohol to do for them, together with how readily they could obtain it and the prevention orientation of their neighborhood, were the dominant correlates of the timing of drinking onset.

### 3.5. Biotyping with *Stacked-Encoders* Embeddings

#### 3.5.1. Biotype Discovery

Unsupervised biotyping of the *Stacked-Encoders* embeddings yielded a stable five-biotype solution (*k* = 5), supported by the joint quality, stability, and survival-separation criteria across *k* = 2-9 (Figure 6B, C) and visualized as well-separated clusters in three-dimensional UMAP (Figure 6A).

**Figure 6.**
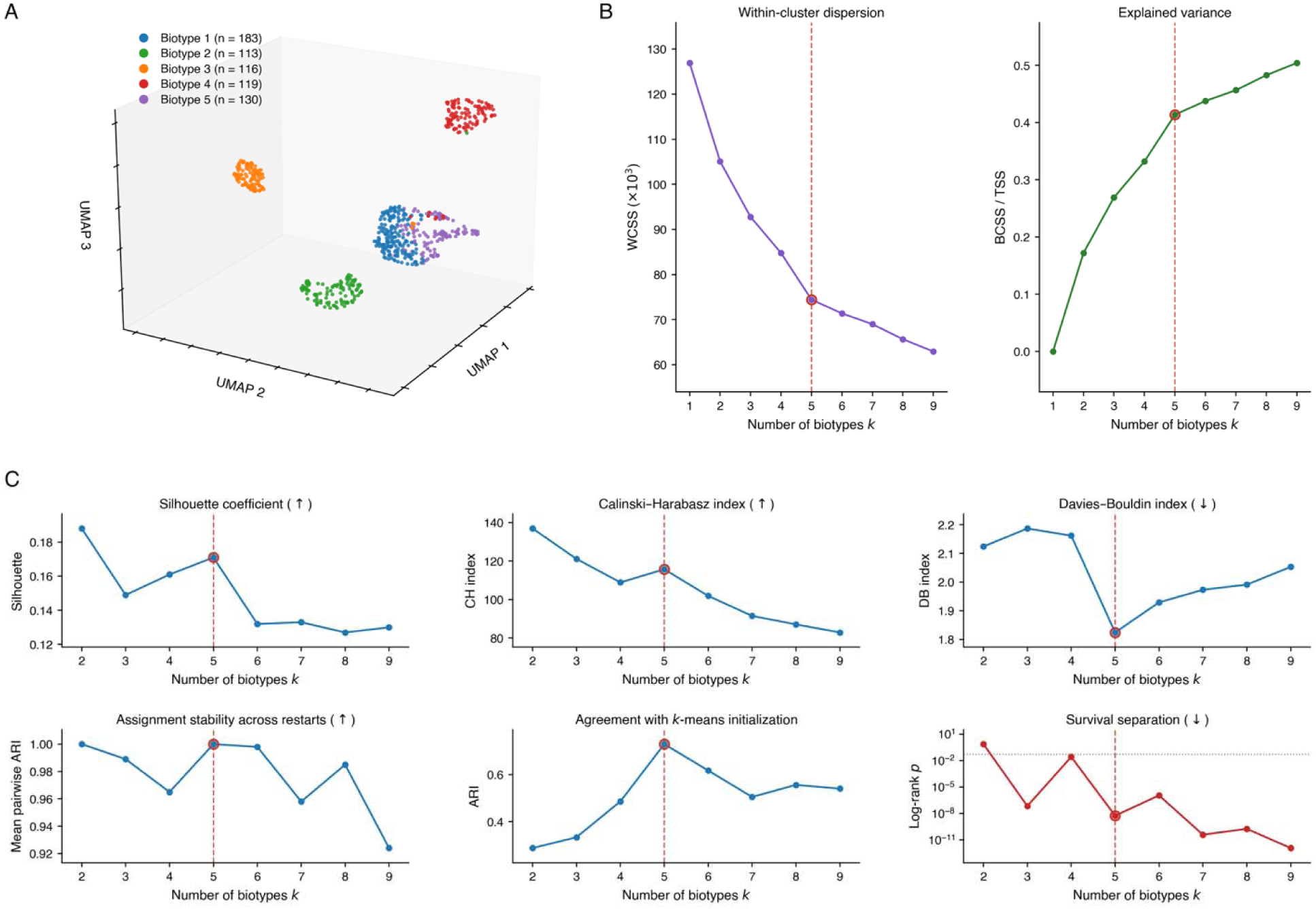
*Stacked-Encoders* embedding biotypes. **A.** Three-dimensional UMAP projection of the 192-dimensional *Stacked-Encoders* embedding, coloured by biotype (k = 5). **B.** Within-cluster dispersion and explained variance across k. **C.** Clustering quality (silhouette, Calinski-Harabasz, Davies-Bouldin), assignment stability (adjusted Rand index), and survival separation (log-rank p) across k = 2-9; the selected solution is k = 5.

#### 3.5.2. Biotype TTE Associations

Time to drinking onset differed across biotypes (Figure 7). In summary, biotype 1 was the latest onset group, while biotype 5 was the earliest. To be specific, mean TTE decreased monotonically from 4.30 ± 1.64 years in biotype 1 through 3.83 ± 1.86 years in biotype 2, 3.55 ± 1.89 years in biotype 3, and 3.42 ± 1.81 years in biotype 4, to 2.86 ± 1.79 years in biotype 5. The omnibus biotype effect was significant both unadjusted (*p* = 1.9 × 10^-10^) and after adjustment for baseline age, sex, and site (*p* = 2.0 × 10^-4^; partial *η*^2^ = 0.033). Two pairwise contrasts survived Holm-Šidák correction: biotype 5 versus biotype 1 (adjusted Δ = -0.86 years, Standard Error (SE) = 0.20, *p* = 0.0002) and biotype 5 versus biotype 2 (adjusted Δ = -0.73 years, SE = 0.21, *p* = 0.006).

**Figure 7.**
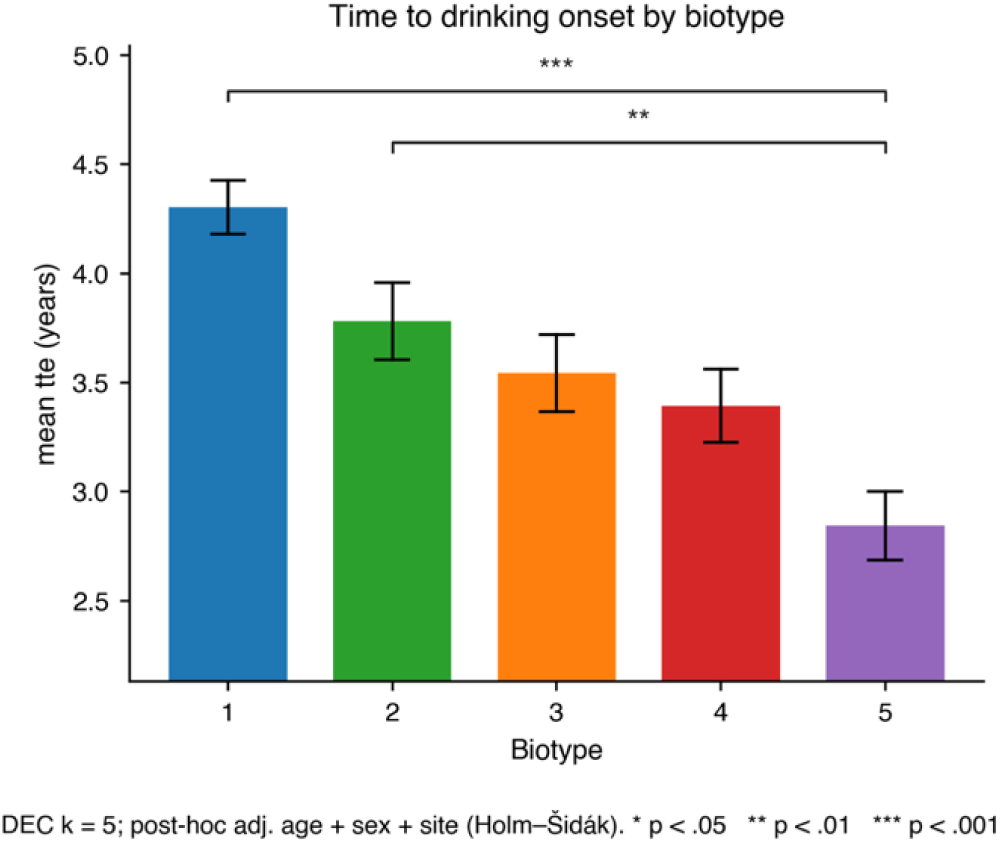
Biotype ANCOVA and post-hoc of TTE differences

#### 3.5.3. Biotype Neuroimaging Associations

The biotypes were further distinguished by neuroimaging phenotypes (Figure 8A-C). In resting-state ReHo, the right hippocampus (F = 3.98, partial η² = 0.024, FDR *p* = 0.027), right amygdala (F = 3.12, FDR *p* = 0.045), and left hippocampus (F = 3.04, FDR *p* = 0.045) differed significantly across biotypes, with post-hoc contrasts localizing the effects to biotype 5 (e.g., right hippocampus biotype 5-4 adjusted mean difference = -0.014, *p* = 1.2 × 10^-4^; right amygdala biotype 5-4 adjusted mean difference = -0.016, *p* = 0.001). In DTI orientation-dispersion, the right anterior thalamus tract showed a significant biotype effect (F = 5.48, partial η² = 0.033, FDR *p* = 0.002), again driven by biotype 5 (biotype 5-2 adjusted difference = -0.012, *p* = 2 × 10^-5^). In FreeSurfer morphometry, a right suborbital sulcs (ventromedial prefrontal) thickness effect reached nominal significance and a significant biotype 5-4 post-hoc contrast (Δ = 0.163, *p* = 0.002) but did not survive whole-map FDR correction. Together, the earliest-onset biotype (biotype 5) was characterized by altered hippocampal/amygdalar regional homogeneity and anterior white-matter organization, linking the *FocalTab*-derived representation to limbic and prefrontal substrates implicated in adolescent alcohol-use risk.

**Figure 8.**
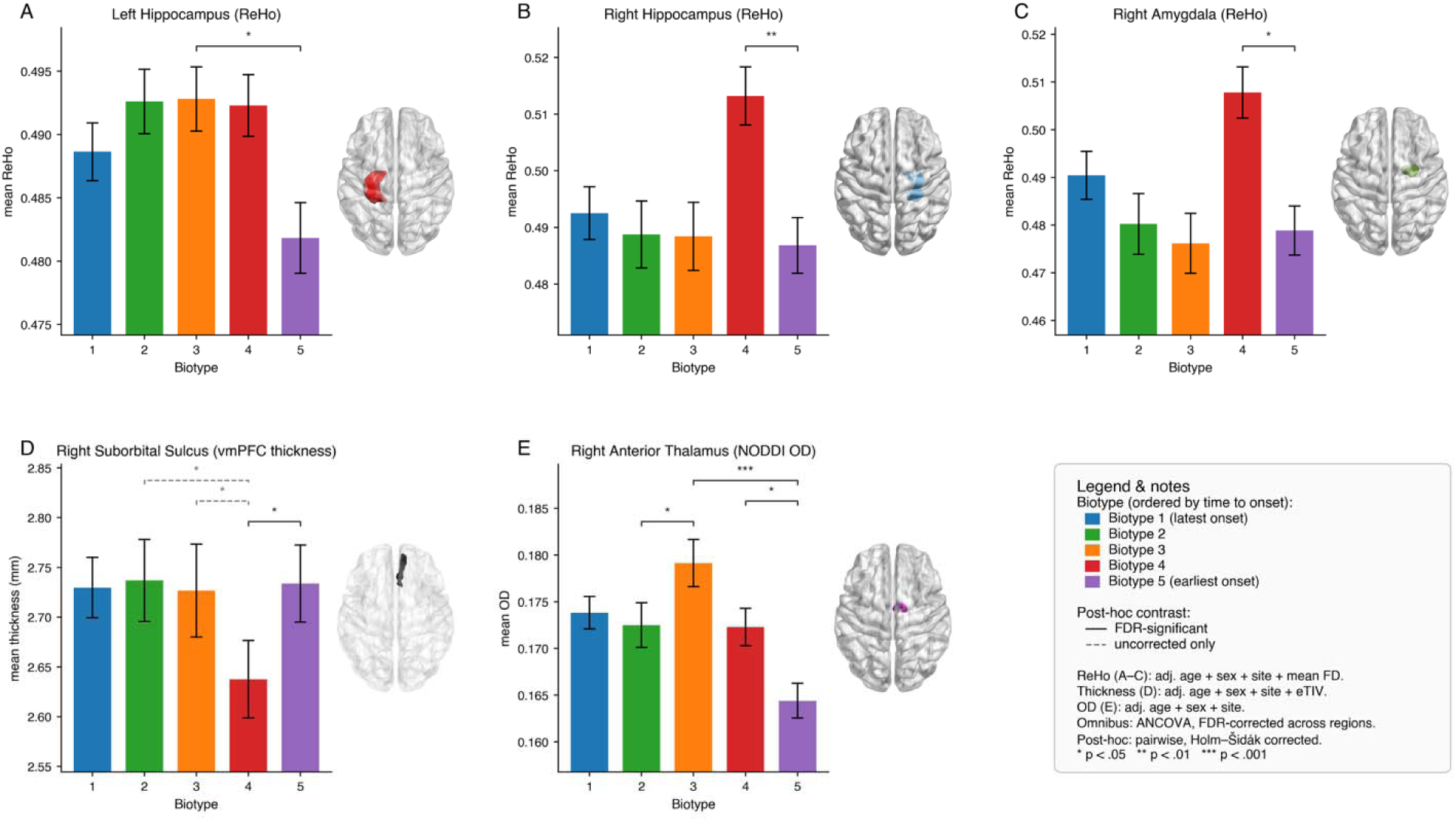
Biotype ANCOVA and post-hoc results (A. ReHo, B. DTI-OD, C. FreeSurfer).

## Discussion

In this study, we developed a stacked-encoder survival framework to predict time from baseline assessment to drinking onset in adolescents using only clinical measures obtained from routine interviews and questionnaires. The framework incorporated 1,582 baseline accessible clinical features spanning behavioral, psychosocial, environmental, and family-history domains, without using neuroimaging inputs. To address the high dimensionality relative to the sample size (n = 661), representation learning was separated from survival modeling: an unsupervised denoising autoencoder and a focal-loss TabPFN (*FocalTab*) encoder were pretrained and subsequently held fixed during survival-head training. Class imbalance between onset events and censored observations was addressed through focal loss during encoder training rather than resampling, thereby preserving the original data distribution.

The Focal Stacked-Encoders representation yielded the strongest overall performance on the held-out test set. Under the random survival forest head, it achieved an Uno C of 0.7654 [0.7020, 0.8286] and a tdAUC of 0.8274 [0.7524, 0.8897]. Under the FCN-CoxPH head, it achieved a Harrell C of 0.7557 [0.6907, 0.8148] and an IBS of 0.1708 [0.1516, 0.1912] (Table 2; Figure 3). These results suggest that the pretrained TabPFN representation can effectively capture predictive structure in high-dimensional tabular data despite the modest sample size (n = 661; p = 1,582), consistent with TabPFN’s design for small tabular datasets through prior-based inference ^18^.

Performance showed a consistent pattern across both survival heads: the stacked representation outperformed the single encoder, which in turn outperformed the raw clinical-feature baseline (Table 2). Relative to the raw-feature baseline (RSF: Uno C = 0.7206, tdAUC = 0.7820, IBS = 0.1809), the stacked representation improved discrimination and time-dependent prediction by approximately 0.045 while reducing the IBS. Each encoder provided incremental value when used alone: the autoencoder achieved an Uno C of 0.7367 and a tdAUC of 0.7930, whereas the *FocalTab* encoder achieved 0.7318 and 0.7747, respectively, compared with 0.7107 and 0.7713 for the corresponding raw-feature baseline under FCN-CoxPH. However, neither matched the combined representation, suggesting that the two encoders capture complementary information. One possible explanation is that the autoencoder reduces sparsity and redundancy in the questionnaire-derived feature space, whereas the pretrained tabular encoder provides a prior over feature relationships that may be difficult to learn from the 462-participant training set alone. More broadly, representation learning has been used in similarly sized studies to reduce the dimensionality of high-dimensional measurements and facilitate downstream statistical or predictive modeling ^11,15,21^.

The choice of fine-tuning objective substantially affected tabular representation quality. The standard cross-entropy TabPFN encoder yielded the weakest performance, in some cases performing below the raw-feature baseline (FCN-CoxPH: Uno C = 0.6885, IBS = 0.2137; RSF: Uno C = 0.6320, tdAUC = 0.6725). In contrast, the focal-loss encoder improved performance over the corresponding baseline (FCN-CoxPH: Uno C = 0.7318), and this advantage persisted after stacking. The focal-loss representation outperformed the cross-entropy representation on discrimination and accuracy for both survival heads (RSF: Uno C = 0.7654 vs. 0.7592, tdAUC = 0.8274 vs. 0.8202; FCN-CoxPH: Uno C = 0.7643 vs. 0.7553), although the autoencoder partially reduced the performance gap. Focal loss ^17^ emphasizes harder-to-classify observations by reducing the contribution of easily classified examples, addressing class imbalance during representation learning without resampling and thus preserving the observed data distribution. This differs from approaches in adolescent substance-use prediction that rely primarily on evaluation metrics ^7^, do not explicitly address low event rates ^16^, or use resampling strategies ^8^. Notably, the RSF showed a larger performance degradation than the Cox head when using the cross-entropy representation, suggesting greater sensitivity to less informative embeddings. Overall, these findings indicate that representation quality had a larger influence on predictive performance than the choice of survival head.

Performance was comparable between female and male participants (Table 3; Figure 4). The Uno C was 0.7374 [0.6234, 0.8355] among females (35 events/44 participants) and 0.7798 [0.6848, 0.8680] among males (40 events/56 participants), with nearly identical time-dependent AUCs (0.8181 vs. 0.8183) and similar IBS (0.1739 vs. 0.1683). The model therefore showed comparable predictive performance across the two groups, although larger held-out samples are needed to determine whether onset timing can be predicted equally well across sexes. Prior work in this cohort has identified sex-specific risk architecture for binary heavy-drinking onset ^7^, highlighting the need for future studies to explicitly evaluate sex differences in the prediction of onset timing.

SHAP analysis of the best-performing model identified four coherent predictor domains among the highest-ranked features (Figure 5): substance access, positive alcohol expectancies, neighborhood and community engagement, and a circadian marker. These findings were broadly consistent with an independent analysis of the same cohort that used a longitudinal deep-learning model and permutation testing rather than baseline prediction with SHAP. That study likewise identified alcohol expectancies, peer drinking, and alcohol access as prominent predictors, while finding no significant contribution from neuroimaging, cognitive, or mental-health measures ^9,10^. Together, these convergent findings highlight the importance of psychosocial and environmental factors in predicting adolescent drinking onset.

Unsupervised clustering of the stacked-encoder embeddings identified five stable biotypes with distinct onset timing (Figure 6). Mean time to onset decreased monotonically from 4.30 ± 1.64 years in biotype 1 to 2.86 ± 1.79 years in biotype 5, with a significant omnibus effect both before adjustment (p = 1.9 × 10^-10^) and after adjustment for baseline age, sex, and site (p = 2.0 × 10^-4^, partial η² = 0.033); two pairwise contrasts remained significant after Holm-Šidák correction. Importantly, the biotypes also differed on phenotypes not used for clustering: regional homogeneity in the bilateral hippocampus and right amygdala and orientation dispersion in the right anterior thalamic region remained significant after FDR correction, with effects concentrated in the earliest-onset group, whereas a ventromedial prefrontal cortical thickness effect was only nominally significant. These findings implicate limbic and thalamo-prefrontal systems previously associated with reward processing and inhibitory control during adolescence ^46,47^.

Several limitations should be acknowledged. First, all analyses were conducted within a single cohort and a single training-validation-test partition, without external validation. Second, subgroup analyses were based on small test strata (44 females and 56 males); therefore, these estimates were interpreted descriptively rather than formally compared. Third, biotypes were derived using all 661 participants, so their robustness was evaluated through restart stability and internal clustering indices rather than out-of-sample assignment. Future studies should evaluate model performance across cross-validation folds and leave-one-site-out partitions, test whether biotype membership provides incremental prognostic value when added to survival models and examine whether neuroimaging phenotypes mediate the association between biotype membership and drinking-onset timing.

## Data Availability

All data produced are available online at https://ncanda.org/datasharing.php.

## Acknowledgement

Research reported in this publication was supported by the National Institute of Alcohol Abuse and Alcoholism of the National Institutes of Health under Award Number R21AA032098, R01 AA029127, P60 AA031124, F32AA032170, L30AA032656, and P50AA030407-5126 Pilot Core grant. This project is also supported by the U.S. National Science Foundation under Award Numbers 2500836 and 2614824, and the National Cancer Institute of the National Institutes of Health under Award Number R03CA317707. This research was supported by the State of Nebraska through the Pediatric Cancer Research Group, part of the Child Health Research Institute (CHRI), as well as the CHRI pilot. The content is solely the responsibility of the authors and does not necessarily represent the official views of the funding organizations.

